# Outcomes of possible and probable rheumatic fever: a cohort study using northern Australian register data, 2013-2019

**DOI:** 10.1101/2023.02.01.23285323

**Authors:** Laura Goddard, Mirjam Kaestli, Enes Makalic, Anna P. Ralph

**Affiliations:** Global and Tropical Health Division, Menzies School of Health Research, Charles Darwin University, Darwin, Northern Territory, Australia; School of Global and Population Health, University of Melbourne, Melbourne, Victoria, Australia; Research Institute for the Environment and Livelihoods, Charles Darwin University, Darwin, Northern Territory, Australia

## Abstract

**Background:** Outcomes after acute rheumatic fever (ARF) diagnosis are variable, ranging from recovery to development of severe rheumatic heart disease (RHD). There is no diagnostic test. Evaluation using the Australian clinical diagnostic criteria can result in a diagnosis of ‘definite’, ‘probable’ or ‘possible’ ARF. The ‘possible’ category was introduced in 2013 in Australia’s Northern Territory (NT). Our aim was to compare longitudinal outcomes after a diagnosis of definite, probable or possible ARF.

**Methods:** We extracted data from the NT RHD register for Indigenous Australians with an initial diagnosis of ARF during the 5.5-year study period (01/01/2013 – 30/06/2019). Descriptive statistics were used to describe the demographic and clinical characteristics at initial ARF diagnosis. Kaplan-Meier curves were used to assess the probability of survival free of disease progression and the cumulative incidence risk at each year since initial diagnosis was calculated. Cox proportional hazards regression was used to determine whether time to disease progression differed according to ARF diagnosis and whether progression was associated with specific predictors at diagnosis. A multinomial logistic regression model was performed to assess whether ARF diagnosis was associated with RHD outcome and to assess associations between ARF diagnosis and clinical manifestations. A generalised linear mixed model (GLMM) was developed to assess any differences in the long-term antibiotic adherence between ARF diagnosis categories and to examine longitudinal trends in adherence.

**Results:** There were 913 initial ARF cases, 732 with normal baseline echocardiography. Of these, 92 (13%) experienced disease progression: definite ARF 61/348 (18%); probable ARF 20/181 (11%); possible ARF 11/203 (5%). The proportion of ARF diagnoses that were uncertain (i.e. possible or probable) increased over time, from 22/78 (28%) in 2013 to 98/193 (51%) in 2018. Cumulative incidence risk of any disease progression at 5.5 years was 33.6 (23.6–46.2) for definite ARF, 13.5 (8.8–20.6) for probable and 11.4% (95% CI 6.0–21.3) for possible ARF. The probability of disease-free survival was lowest for definite ARF and highest for possible ARF (p=0.004). Cox proportional hazards regression indicated that disease progression was 2.19 times more likely in those with definite ARF than those with possible ARF (p=0.026). Progression to RHD was reported in 37/348 (11%) definite ARF, 10/181 (6%) probable ARF, and 5/203 (2%) possible ARF. The multinomial logistic regression model demonstrated a significantly higher risk of progression from no RHD to RHD if the initial diagnosis was definite compared to possible ARF (p<0.001 for both mild and moderate-severe RHD outcomes). The GLMM estimated that patients with definite ARF had a significantly higher adherence to antibiotic prophylaxis compared with probable ARF (p=0.024).

**Conclusion:** These data indicate that the ARF diagnostic categories are being applied appropriately, are capturing more uncertain cases over time, provide a useful way to stratify risk and guide prognosis, and can help inform practice. Possible ARF is not entirely benign; some cases progress to RHD.

## Introduction

In Australia, high burdens of acute rheumatic fever (ARF) and rheumatic heart disease (RHD) occur among Aboriginal and Torres Strait Islander peoples, particularly across northern and central Australia.(1) There is no diagnostic test for ARF. This impairs case detection and opportunities for early intervention with antibiotic secondary prophylaxis.(2) The Jones Criteria are syndromic criteria to diagnose ARF, dividing the clinical features into major and minor manifestations based on their prevalence and specificity.(3) The Jones Criteria have been regularly revised in response to evolving clinical knowledge and epidemiology, to reduce overdiagnosis in high-resource settings where ARF has become uncommon.(4-6) As a result, they became less sensitive for detection of cases in high-incidence populations. Different considerations were therefore added for high-risk groups (3, 5, 6), but to further avoid missed diagnoses in high-prevalence Aboriginal communities in Australia, additional diagnostic categories of ‘possible’ and ‘probable’ ARF were proposed.(7) This proposal was adopted and since 2013, diagnoses in Australia have been grouped as: possible ARF, probable ARF, definite ARF, and RHD. The distinction between probable and possible is subjective and requires thoughtful clinical decision making. These categories were further clarified in the revised 2020 Australian Guidelines (Box 1).(3) This is in contrast to international guidelines that only articulate two categories: definite and probable ARF.(6)

### Box 1

**Acute rheumatic fever diagnostic categories, 2020 Australian Guidelines** (6)

**Definite ARF**

acute presentation which fulfils the 2015 Revised Jones diagnostic criteria for ARF.

**Probable ARF**

acute presentation which does not fulfil Jones diagnostic criteria for ARF, missing one major or one minor criterion or lacking evidence of preceding streptococcal infection, but ARF is still considered the most likely diagnosis.

**Possible ARF**

acute presentation which does not fulfil Jones diagnostic criteria for ARF, missing one major or one minor criterion or lacking evidence of preceding streptococcal infection, and ARF is considered uncertain but cannot be ruled out.

Previous research has described progression from ARF to RHD and other outcomes such as morbidity and mortality rates.(8, 9) In Australia’s Northern Territory (NT), the risk of progression from definite ARF to RHD is high, with a cumulative incidence of progression of 27.1% at 1 year after diagnosis, 44.0% at 5 years and 51.9% at 10 years.(9) Adherence to penicillin to prevent rheumatic fever recurrences reduces risk, with a dose-dependent effect.(10) Previous studies routinely exclude possible ARF and often also exclude probable ARF cases, so long-term outcomes of these diagnoses remain unknown.

To provide patients with better prognostic information, determine whether clinicians appear to be applying diagnostic categories appropriately, and reflect on whether current management guidelines (3) are appropriate, the aim of this study is to describe patient outcomes after a diagnosis of possible or probable (collectively, ‘uncertain’) ARF, compared with definite ARF. Further aims are to determine whether uncertain diagnoses differ from definite ARF in demographic or clinical characteristics, severity of RHD if there is progression to RHD, and penicillin adherence over time.

## Methods

### Study design and data sources

We used a retrospective cohort design and Cox proportional hazards regression to analyse the probability of disease progression after an initial diagnosis of ARF. Sub-analyses on initial ARF diagnoses with RHD outcome and adherence to penicillin adherence were conducted using multinomial logistic regression and generalised linear mixed-effects models (GLMM) respectively. Data were extracted from the NT RHD Register (the Register) for a 5.5-year period (01/01/2013 – 30/06/2019). The Register was established in 1997 as a tool to support and coordinate care. Since then, all notified cases of ARF (including recurrent episodes) and RHD in the NT have been recorded, capturing approximately 3,500 individuals at the time of data extraction. The Register includes data on patient demographics, clinical features of ARF and RHD, medical appointments and secondary penicillin prophylaxis, which is generally required long-term after probable and definite ARF, and short term (12 months) after possible ARF. Mortality data from the NT Births, Deaths and Marriages database are reviewed monthly and integrated with the Register.

### Study population and sample

The study population was defined as Indigenous Australians residing in the NT, which is approximately 74,546 or 30% of the total NT population.(11, 12) The Register contained information on 1,140 individuals with 1,567 diagnoses during the study period. Indigenous Australians residing in the NT who were registered with an initial diagnosis of possible, probable or definite ARF on the Register during the study period were included. Exclusions comprised non-Indigenous people (11; 1%); cases whose first documented ARF episode was labelled a recurrence (216; 19%); and concurrent ARF and RHD diagnoses (180; 16%). Those whose first documented episode was labelled a recurrence were assumed to have had a previous diagnosis of ARF before the study period began or in a different jurisdiction. One case whose initial diagnosis date was on the final day of the study period was excluded from the disease progression analyses due to a survival time of zero. Concurrent ARF and RHD diagnoses (180 cases) were similarly excluded from disease progression analyses since they had a disease-free survival time of zero. These cases represent severe ARF which has already progressed to RHD at the time of first echocardiogram, or ARF occurring in someone with previously unrecognised latent RHD, or documentation of rheumatic carditis in the register as RHD (since distinguishing rheumatic carditis from established RHD is not always possible, and misclassification may occur during clinical reporting or data entry). The exclusions mean that the most severe form of ARF (definite ARF with carditis classified as RHD, or carditis complicating RHD), were excluded from the Cox regressions but were still accounted for in the multinomial model. The final sample size for the Cox regression was 732 cases and 913 cases for the multinomial model.

### Outcomes

The main outcome of interest was time to disease progression. Disease progression was defined as progression from possible to probable ARF, definite ARF or RHD; probable to definite ARF or RHD; or definite ARF to definite ARF recurrence or RHD at any time during the study period (Table 1). Individuals could progress and regress more than once during the study period, however only the first progression was counted.

**Table 1.** Definitions of disease progression.

| Initial ARF diagnosis | Subsequent diagnosis |
| --- | --- |
| ARF possible | ARF probable, ARF definite or RHD |
| ARF probable | ARF definite or RHD |
| ARF definite | ARF definite recurrence or RHD |

We also calculated the proportion of those without baseline carditis who progressed to RHD and the risk to develop mild or moderate to severe RHD compared to no RHD based on initial ARF diagnosis.

Adherence to secondary prophylaxis with benzathine benzylpenicillin (BPG) was calculated as the percentage of received doses / prescribed doses x100. BPG is usually required once every 28 days for ARF secondary prophylaxis, i.e. 13 doses per year, but may be prescribed every 21 days, i.e. 17 doses per year, in more severe or breakthrough cases. The number of prescribed doses was estimated by dividing the number of days that BPG was prescribed (last dose date minus first dose date) divided by 21 or 28 for 3-weekly and 4-weekly BPG respectively. Adherence calculations were restricted to cases that had been prescribed at least 6 doses of BPG (equivalent to 168 days for a 4-weekly regimen). Adherence was considered good if ≥80% of injections were received.

### Predictors

Predictors used in the analyses included initial ARF diagnosis, sex, age group, clinical manifestations, and adherence to secondary prophylaxis. Initial ARF diagnosis was defined as the first diagnosis of possible, probable or definite ARF on the Register. Sex was classified as per the NT RHD register (male, female, unknown). Age at initial diagnosis was grouped as 0-4, 5-14, 15-24, ≥25 years; these have different risk profiles for ARF and RHD.(3) Clinical manifestations were grouped as major manifestations (carditis; arthritis or ‘joint’; Sydenham’s Chorea; and subcutaneous nodules and erythema marginatum or ‘skin’), minor manifestations (fever; elevated ESR or CRP; prolonged P-R interval on ECG), and evidence of Group A streptococcal infection (elevated serological titre or cultured from throat swab) as per the RHDAustralia Clinical Guidelines.(3) The definition of adherence is described in the outcomes section. Individuals who had missing data for adherence are excluded from analyses as these data are unlikely to be missing at random and keeping these individuals in the analysis may therefore introduce bias.

### Statistical analysis

Descriptive statistics were used to describe the demographic and clinical characteristics at initial ARF diagnosis (possible, probable, definite). Kaplan-Meier curves and log rank test were used to assess the probability of survival free of disease progression and the cumulative incidence risk at each year since initial diagnosis was calculated. Cox proportional hazards regression was used to determine whether time to disease progression differed according to ARF diagnosis category (definite, probable, possible), and whether it was predicted by age group at diagnosis, sex, adherence to secondary prophylaxis or clinical manifestations (joint manifestation-only and joint manifestation with carditis and/or chorea) at diagnosis. Hazard ratios, 95% CI and p-values are reported.

A multinomial logistic regression model was performed to assess whether ARF diagnosis type (definite, probable, possible) was associated with RHD outcome (no RHD, mild RHD moderate-severe RHD). Moderate and severe RHD were combined to increase observations per group level. Predictors included age group, sex, drug adherence and clinical manifestations (carditis-only or joint manifestations-only. Chorea-only was excluded due to the small number of observations). Multinomial logistic regression was also used to assess associations between clinical manifestations (Jones major criteria) and ARF diagnosis. Relative risk ratios, 95% confidence intervals (CI) and p-values were calculated for clinical manifestations, age at diagnosis and sex, using definite ARF as the reference group.

A GLMM (beta regression family) was developed to assess any differences in the long-term antibiotic adherence between ARF diagnosis categories and examine longitudinal trends in adherence (glmmTMB package in R). This took into account the number of years receiving BPG prophylaxis and a random intercept for patient identification. Model residuals were checked for lack of patterns across fitted values and predictors (DHARMa package) and no temporal autocorrelation.

Data were analysed using STATA version 15.1 (STATA Corp, College Station, TX), R (v4.1.3; R Project for Statistical Computing, Vienna, Austria) and Microsoft Excel (2016).

### Ethics approval

Study approval was granted by the Human Research Ethics Committee of the Northern Territory Department of Health and Menzies School of Health Research (2019-3482). This approval was registered with the University of Melbourne’s Medicine and Dentistry Human Ethics Sub-Committee (1955194). Access to register data was approved by the data custodian, Top End Health Service, NT Department of Health.

## Results

### Demographic and clinical characteristics

During the study period there were 913 initial diagnoses of ARF among eligible register participants: 509 (56%) definite, 196 (21%) probable and 208 (23%) possible ARF. All ARF types were more common among females than males and in those aged 5-14 years compared with other age groups (**Error! Reference source not found**.).

Joint pain was the most common major Jones criterion for all ARF diagnostic categories. Evidence of streptococcal infection (considered essential) was missing in 2% of cases classified as definite ARF, and in approximately 10% of possible and probable ARF cases. There were 2 deaths during the study period. The proportion of those adhering to penicillin secondary prophylaxis was similar for possible and probable ARF but higher for definite ARF (Table 2): the proportion receiving ≥80% of scheduled BPG injections was 37% and 46% among possible and probable cases respectively, compared with 56% among definite ARF cases. However, the proportion of possible ARF cases with unknown adherence was double that among probable ARF and definite ARF cases (**Error! Reference source not found**.). Each year, the number of ARF diagnoses increased (Fig 1).

**Table 2.** Characteristics at initial acute rheumatic fever diagnosis among Indigenous Australians, NT, 2013-2019

| Characteristic | ARF |  |  |  |  |  | Total |  |
| --- | --- | --- | --- | --- | --- | --- | --- | --- |
|  | Possible |  | Probable |  | Definite |  |  |  |
|  | n | (%) | n | (%) | n | (%) | n | (%) |
| Cases | 208 | (100) | 196 | (100) | 509 | (100) | 913 | (100) |
| Sex |  |  |  |  |  |  |  |  |
| Male | 97 | (47) | 87 | (44) | 243 | (48) | 427 | (47) |
| Female | 111 | (53) | 109 | (56) | 266 | (52) | 486 | (53) |
| Age group (years) |  |  |  |  |  |  |  |  |
| 0-4 | 25 | (12) | 11 | (6) | 27 | (5) | 63 | (7) |
| 5-14 | 113 | (54) | 92 | (47) | 298 | (59) | 503 | (55) |
| 15-24 | 37 | (18) | 53 | (27) | 107 | (21) | 197 | (22) |
| ≥25 | 33 | (16) | 40 | (20) | 77 | (15) | 150 | (16) |
| Median, IQR | 10 | 6-19 | 14 | 9-22 | 12 | 9-18 | 15 | 8-19 |
| Year of diagnosis |  |  |  |  |  |  |  |  |
| 2013 | 9 | (4) | 13 | (7) | 56 | (11) | 78 | (9) |
| 2014 | 13 | (6) | 11 | (6) | 72 | (14) | 96 | (11) |
| 2015 | 18 | (9) | 28 | (14) | 77 | (15) | 123 | (13) |
| 2016 | 40 | (19) | 36 | (18) | 81 | (16) | 157 | (17) |
| 2017 | 38 | (18) | 52 | (27) | 87 | (17) | 177 | (19) |
| 2018 | 58 | (28) | 40 | (20) | 95 | (19) | 193 | (21) |
| 2019* | 32 | (15) | 16 | (8) | 41 | (8) | 89 | (10) |
| Adherence (%) |  |  |  |  |  |  |  |  |
| <50 | 16 | (7) | 22 | (11) | 52 | (10) | 90 | (10) |
| 50-79 | 47 | (23) | 57 | (29) | 120 | (24) | 224 | (24) |
| 80-99 | 47 | (23) | 55 | (28) | 164 | (32) | 266 | (29) |
| ≥100 | 36 | (17) | 36 | (19) | 109 | (21) | 181 | (20) |
| Unknown † | 62 | (30) | 26 | (13) | 64 | (13) | 152 | (17) |
| Clinical manifestation ‡ |  |  |  |  |  |  |  |  |
| Major |  |  |  |  |  |  |  |  |
| Carditis | 6 | (3) | 14 | (7) | 167 | (33) | 187 | (20) |
| Joint | 172 | (83) | 182 | (93) | 433 | (85) | 787 | (86) |
| Sydenham chorea | 3 | (1) | 0 | (0) | 62 | (12) | 65 | (7) |
| Skin | 2 | (1) | 1 | (1) | 7 | (1) | 10 | (1) |
| Minor |  |  |  |  |  |  |  |  |
| Fever | 51 | (24) | 60 | (31) | 340 | (67) | 451 | (49) |
| Elevated ESR or CRP | 12 | (14) | 9 | (6) | 11 | (2) | 32 | (5) |
| Prolonged P-R interval on ECG | 10 | (5) | 19 | (10) | 196 | (39) | 225 | (25) |
| Other |  |  |  |  |  |  |  |  |
| Elevated GAS serological titre | 183 | (88) | 168 | (86) | 480 | (94) | 831 | (91) |
| GAS cultured from throat swab | 5 | (2) | 7 | (4) | 18 | (4) | 30 | (3) |
| Not recorded | 6 | (3) | 1 | (1) | 0 | (0) | 7 | (1) |

ARF = acute rheumatic fever; GAS = group A beta haemolytic streptococcus; ESR = erythrocyte sedimentation rate; CRP = C-reactive protein; ECG = electrocardiogram; CM = clinical manifestation
\* Note: only 6 months of data
† Note: includes those where prophylaxis data is not recorded and where prophylaxis was prescribed for <6 months.
‡ Note: cases may have more than one clinical manifestation, therefore the total does not add up to 100%
¥ Note: Elevated GAS serological titre or GAS cultured from throat swab only

**Fig 1:**
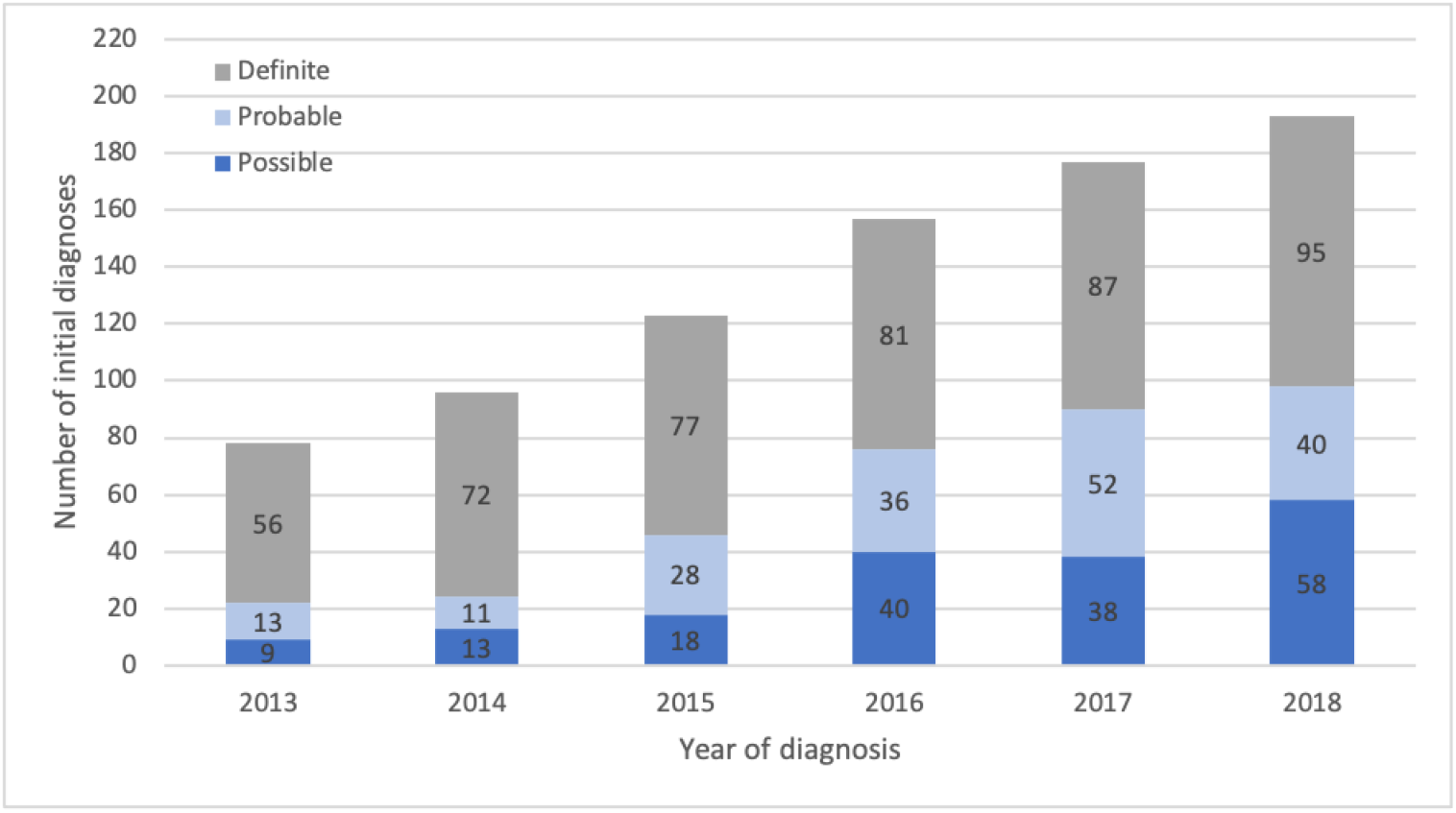
Number of ARF diagnoses among Indigenous Australians, Northern Territory, 2013-2018. ARF indicates acute rheumatic fever.

The proportion that was uncertain increased from 22/78 (28%) in 2013 when these diagnostic categories started to be recorded, to 98/193 (51%) in 2018 (**Error! Reference source not found**.).

### Disease progression

After excluding cases with zero disease-free survival time, a total of 92 (13%) cases experienced disease progression during the study period. Of those with an initial diagnosis of definite ARF, 61/348 (18%) experienced disease progression, compared to 20/181 (11%) with probable ARF and 11/203 (5%) with possible ARF. Of those who progressed, 37/61 (61%) with definite ARF, 10/20 (50%) with probable ARF, and 5/11 (45%) with possible ARF (total of 52/92 or 57%) were diagnosed with RHD during the study period. Overall, progression to RHD was reported in 37/348 (11%) definite ARF, 10/181 (6%) probable ARF, and 5/203 (2%) possible ARF.

Possible ARF had the best probability of survival free of disease progression and definite ARF had the worst survival probability (*p*=0.0043; log rank test; Fig 2).

**Fig 2:**
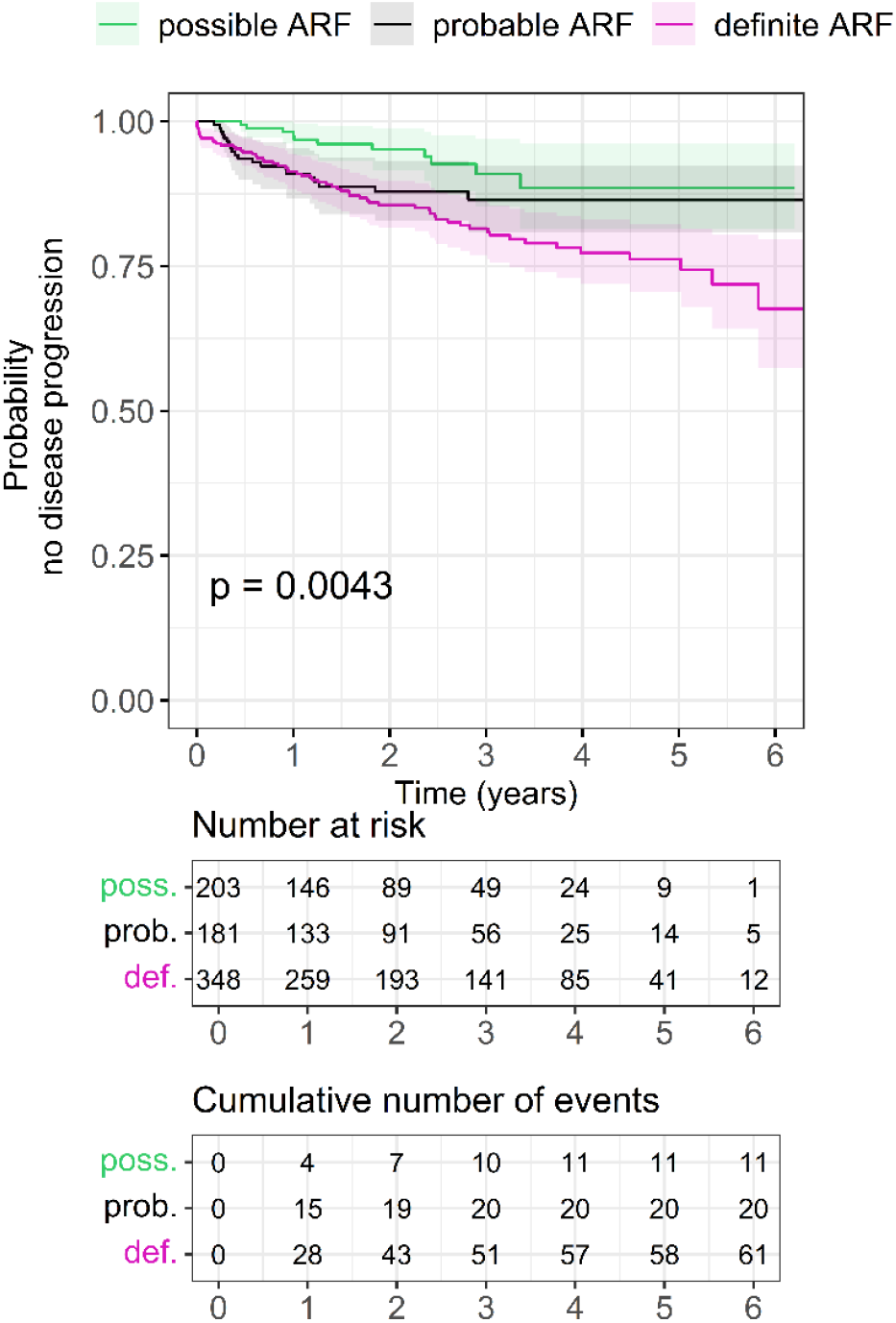
Kaplan-Meier progression-free survival estimates by initial ARF diagnosis among Indigenous Australians, Northern Territory, 2013-2019. ARF = acute rheumatic fever.

For possible and probable ARF, the risk of disease progression continued to increase each year up to 4 years and 3 years since initial diagnosis respectively. For definite ARF, the risk of disease progression continued to increase each year of the follow-up period (Table 3).

**Table 3:** Cumulative incidence risk of disease progression by year of follow-up among Indigenous Australians, Northern Territory, 2013-2019.

| Year | Possible ARF<br>%, CI (95%) | Probable ARF<br>%, CI (95%) | Definite ARF<br>%, CI (95%) |
| --- | --- | --- | --- |
| 1 | 2.46 (0.93 – 6.44) | 9.03 (5.54 – 14.55) | 8.66 (6.05 – 12.32) |
| 2 | 4.85 (2.31 – 10.03) | 12.13 (7.87 – 18.45) | 14.43 (10.87 – 19.02) |
| 3 | 9.05 (4.72 – 17.00) | 13.54 (8.78 – 20.59) | 17.98 (13.83 – 23.19) |
| 4 | 11.44 (5.97 – 21.33) | 13.54 (8.78 – 20.59) | 22.19 (17.22 – 28.33) |
| 5 | 11.44 (5.97 – 21.33) | 13.54 (8.78 – 20.59) | 23.26 (18.01 – 29.73) |
| 5.5* | 11.44 (5.97 – 21.33) | 13.54 (8.78 – 20.59) | 33.55 (23.61 – 46.21) |

\*Only six months of data are available for the final year

Cox proportional hazards regression indicated that disease progression was 2.19 times more likely in those with definite ARF than those with possible ARF (p*=*0.026) (Table 4).

**Table 4.** Cox proportional hazards analysis of predictors associated with disease progression among Indigenous Australians, Northern Territory, 2013-2019.

| Variable | Hazard Ratio | Confidence Interval (95%) | P value |
| --- | --- | --- | --- |
| **ARF Diagnosis** |  |  |  |
| Possible ARF | Ref |  |  |
| Probable ARF | 1.44 | 0.66 – 3.15 | 0.353 |
| Definite ARF | 2.19 | 1.10 – 4.34 | 0.026 |
| **Sex** |  |  |  |
| Male | Ref |  |  |
| Female | 1.06 | 0.67 – 1.67 | 0.811 |
| **Age group (years)** |  |  |  |
| 0-4 | 0.73 | 0.26 – 2.06 | 0.552 |
| 5-14 | Ref |  |  |
| 15-24 | 1.28 | 0.75 – 2.20 | 0.366 |
| ≥25 | 1.71 | 0.93 – 3.15 | 0.084 |
| **Adherence** |  |  |  |
| <50% | 0.31 | 0.09 – 1.11 | 0.072 |
| 50-79% | 1.48 | 0.66 – 3.29 | 0.337 |
| 80-99% | Ref |  |  |
| ≥100% | 2.22 | 0.86 – 5.78 | 0.100 |
| **Clinical manifestations** |  |  |  |
| Carditis and/or chorea with or without joint manifestation | Ref |  |  |
| Joint manifestation only (i.e. no carditis, no chorea) | 0.48 | 0.24 – 0.95 | 0.035 |

ARF = acute rheumatic fever
1. Sample size for this analysis is 595 cases (137 cases missing drug adherence data and 181 cases with zero survival time were excluded) 2. Proportional hazard assumptions were not met for drug adherence nor joint manifestation and therefore, an interaction term with linear time was fitted for these 2 factors (likelihood ratio test $\text{Chi}^2(3) = 14.9$ ; $\text{Chi}^2(1) = 7.4$ , $P < 0.01$ for both). The time varying effect for joint manifestation was 1.7 ( $P = 0.012$ ) and 2.0 for prophylaxis adherence of <50% ( $P = 0.010$ ), both indicating a weakening protective effect with time.

The analysis also indicated that having joint manifestations only with no carditis nor chorea at initial diagnosis of ARF was protective, with this group being 52% less likely to experience disease progression compared with those who had other clinical manifestations including carditis and/or chorea at initial diagnosis and with or without joint manifestation (p=0.035) (Table 4). Disease progression was equally likely between sexes and age groups. In this dataset, the association between penicillin adherence and progression was counterintuitive and not statistically significant. People who had higher adherence were those who had disease progression (Table 4).

### Risk of developing rheumatic heart disease

The proportion of those who had no baseline carditis but still progressed to RHD is presented by clinical manifestation (joint-only or chorea-only) in Table 5. Of those who progressed to RHD with joint-only manifestations, 46% (17/37) progressed after one year. Of those who progressed to RHD with chorea-only manifestations, only 3% (1/30) progressed after one year. An additional case with neither chorea nor joint manifestations also progressed to RHD after one year, bringing the total cases with progression to RHD after one year to be 19 cases.

**Table 5:**
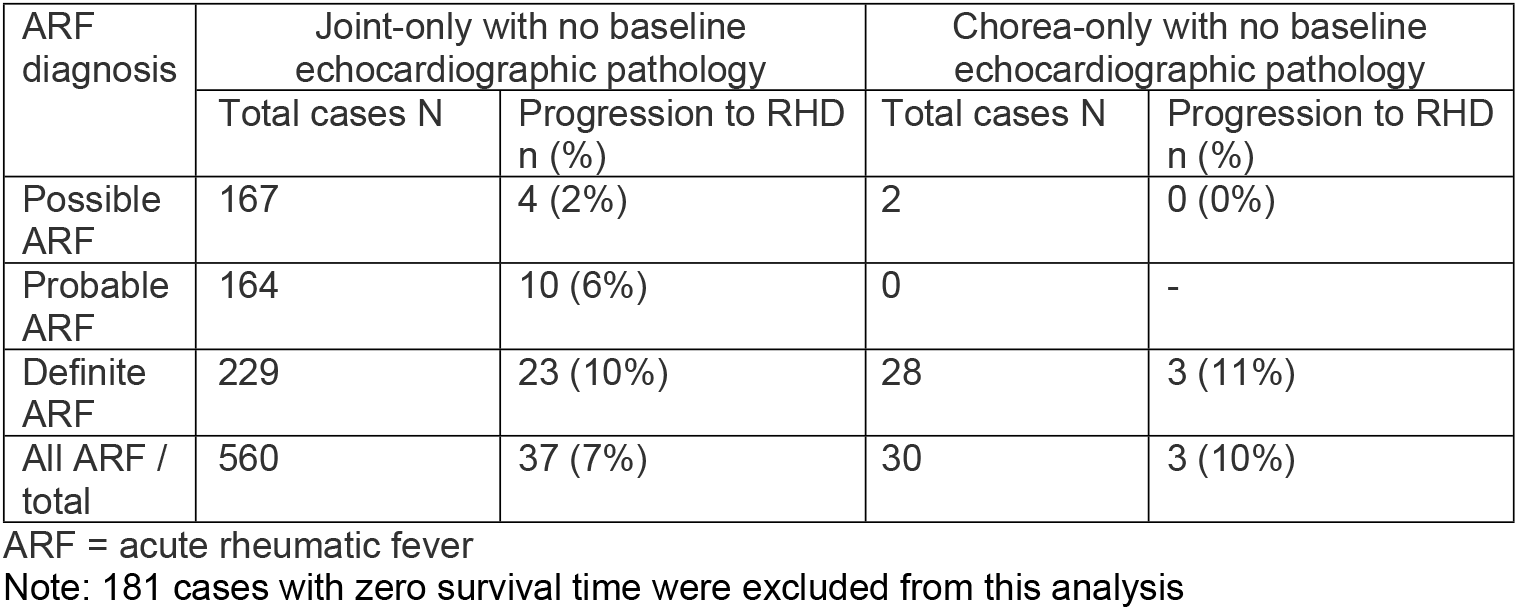
Progression to RHD by clinical manifestation among Indigenous Australians, Northern Territory, 2013-2019.

The multinomial logistic regression model demonstrated a significantly higher risk of progression from no RHD to RHD if the initial diagnosis was definite compared to possible ARF (p<0.001 for both mild and moderate-severe RHD outcomes) (Table 6).

**Table 6:** Multinomial logistic regression of initial ARF diagnosis and development of RHD, by RHD severity, among Indigenous Australians, Northern Territory, 2013-2019.

| Multinomial model | Relative Risk Ratio | Confidence Interval (95%) | P value |
| --- | --- | --- | --- |
| Risk to develop mild RHD compared to no RHD after an initial ARF diagnosis |  |  |  |
| **ARF diagnosis** |  |  |  |
| ARF possible | Ref |  |  |
| ARF probable | 2.27 | 0.91-5.64 | 0.078 |
| ARF definite | 6.39 | 2.85-14.35 | < 0.001 *** |
| **Age group** |  |  |  |
| <= 4 yrs | 0.29 | 0.07-1.26 | 0.098 |
| 5-14 yrs | Ref |  |  |
| 15-24 yrs | 1.71 | 1.04-2.81 | 0.034 * |
| >=25 yrs | 1.98 | 1.11-3.54 | 0.021 * |
| **Clinical Manifestations** |  |  |  |
| Carditis only – Yes | 0.95 | 0.27-3.32 | 0.939 |
| Joint only – Yes | 0.49 | 0.32-0.76 | 0.002 ** |
| **Adherence** |  |  |  |
| <50% | 0.49 | 0.24-1.01 | 0.054 |
| 50-79% | 0.77 | 0.47-1.26 | 0.294 |
| 80-99% | Ref |  | 0.591 |
| ≥100% | 0.86 | 0.50-1.48 |  |
| **Gender – Female** |  |  | 0.084 |
| Risk to develop moderate to severe RHD compared to no RHD after an initial ARF diagnosis |  |  |  |
| **ARF diagnosis** |  |  |  |
| ARF possible | Ref |  |  |
| ARF probable | 2.51 | 0.46-13.72 | 0.289 |
| ARF definite | 13.92 | 3.22-60.16 | <0.001*** |
| **Age group** |  |  |  |
| <= 4 yrs | 1.80 | 0.64-5.04 | 0.261 |
| 5-14 yrs | Ref |  |  |
| 15-24 yrs | 1.88 | 0.98-3.61 | 0.059 |
| >=25 yrs | 2.11 | 0.94-4.72 | 0.071 |
| **Clinical Manifestations** |  |  |  |
| Carditis only – Yes | 3.14 | 1.11-8.82 | 0.030* |
| Joint only – Yes | 0.19 | 0.11-0.34 | <0.001*** |
| **Drug adherence** |  |  |  |
| <50% | 0.20 | 0.06-0.74 | 0.015* |
| 50-79% | 0.67 | 0.34-1.31 | 0.237 |
| 80-99% | Ref |  |  |
| ≥100% | 1.41 | 0.75-2.66 | 0.281 |
| **Gender – Female** | 0.90 | 0.52-1.55 | 0.710 |

ARF = acute rheumatic fever; ESR = erythrocyte sedimentation rate; CRP = C-reactive protein; ECG = electrocardiograph; GAS = Group A Streptococcus; \*\*\* P<0.001; \*\* P<0.010; \* P<0.050 Note: sample size for this analysis is 761 cases (151 cases missing drug adherence data were excluded)

There was a strong protective effect for those with joint manifestations only, with a 51% reduced risk to progress to mild RHD and an 81% reduced risk to progress to moderate or severe RHD from no RHD, with all other covariates held constant (p<0.001 for both; Table 6). There was a three-fold increased risk of developing moderate-severe RHD compared to no RHD for patients with carditis-only clinical manifestation (p=0.030).

### Penicillin adherence

The GLMM estimated that patients with definite ARF had a significantly higher adherence to antibiotic prophylaxis compared with probable ARF (p=0.024). Possible ARF, requiring only 12 months of treatment, was excluded from adherence comparisons. Time on prophylaxis was also significantly inversely associated with adherence, with adherence being higher in the first year after diagnosis and falling in years 2 to 6 after ARF diagnosis, for both definite and probable ARF (p<0.001 for all) (Fig 3).

**Fig 3:**
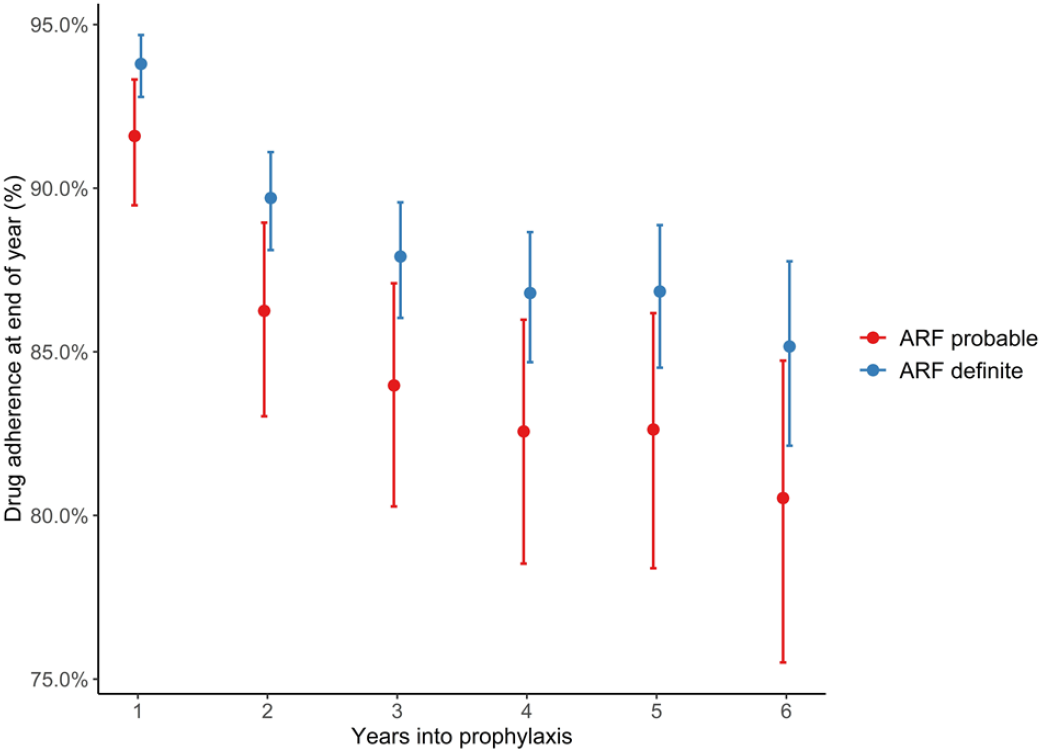
Drug adherence by initial ARF diagnosis and year since diagnosis among Indigenous Australians, Northern Territory, 2013-2019. ARF = acute rheumatic fever.

## Discussion

To our knowledge, this is the first time that outcomes of possible and probable ARF have been determined. We show that since introduction of these diagnostic categories, they now make up over 40% of all initial ARF diagnoses in the Australian jurisdiction with the highest ARF burden. The diagnostic categories appear to be well applied by clinicians: the vast majority of children and young people diagnosed with possible ARF (95%) and probable ARF (89%) did not progress to definite ARF or RHD. These data are reassuring both for clinicians about the appropriate application of these diagnostic categories, and for patients and their families about the likelihood of a good outcome. However, the fact that some patients do progress emphasises that the Revised Jones criteria can still be inadequately sensitive in a high-burden environment, and the categories of ‘possible’ and ‘probable’ ARF add value by capturing some true cases who require institution of secondary prevention and appropriate clinical follow up.

Our findings highlight that definite, probable and possible ARF represent three distinct diagnostic categories with different prognoses. In a high-incidence setting, including the category of ‘possible’ ARF captures an important patient group who might otherwise receive an alternative generic diagnosis such as ‘viral illness’ or ‘musculoskeletal pain’. International guidelines currently only articulate two categories: definite and probable ARF.(6) While many people labelled with possible ARF in our dataset may never have had true ARF, is not entirely benign; 4/200 (2%) with an initial diagnosis of possible ARF and a normal echocardiogram developed RHD; 3 within 12 months (1.5%), and 1 within 24 months (0.5%). The inability to predict which individuals are the ones most likely to progress highlights the need for biomarkers to diagnose and risk-stratify ARF. More research in this domain is needed.(13) Current Australian guidelines recommend 12 months of secondary prophylaxis after a diagnosis of possible ARF.(3) Our findings support this approach, since development of RHD more than 12 months after ‘possible ARF’ appears to be no more common than the background population RHD risk, based on RHD population prevalence of 2% in 5-9 year old children undergoing echocardiographic screening, and 6.8% in 10-15 year olds, in a high-burden NT setting.(14)

Healthcare providers appear to be increasingly confident in accurately applying ‘probable’ and ‘possible’ ARF diagnoses since 2013 when these were defined and introduced into the NT register. When total ARF diagnoses are captured, our data show that the increases over time chiefly represent improving detection of uncertain cases from 22/78 (28%) in 2013 to 98/193 (51%) in 2018, with a smaller proportional increase in numbers of definite ARF (Fig 1). Efforts to improve detection have included promotion of accessible clinical practice guidelines (2, 3, 15) and a diagnosis calculator available for smartphone use.(16)

The clinical manifestations of ARF we report are supported by previous NT data,(17) showing arthritis to be most common major criterion followed by carditis then chorea, with almost all additionally having fever.(18) Rheumatic carditis detected on echocardiogram is diagnostic of definite ARF and should not be classified as possible or probable ARF. Possible and probable cases noted to have echocardiographic valvar disease may have had known RHD and presented with an acute illness not meeting criteria for definite ARF. When a child with RHD presents with fever or elevated inflammatory markers, distinguishing ARF from other febrile conditions with underlying RHD can be difficult and may result in a ‘possible’ or ‘probable’ ARF diagnosis; the child is already prescribed penicillin prophylaxis but a new ARF diagnosis may alter the duration of prophylaxis depending on their age. Sydenham chorea is synonymous with ARF; once alternative causes of chorea have been excluded, a child with chorea (especially if from a high-risk setting such as the NT) receives a diagnosis of definite ARF. The possible and probable cases in this study noted to have chorea may, for instance, have had a transient movement disorder not clearly evident as Sydenham chorea. Definite ARF diagnoses require demonstration of recent streptococcal infection, except for when Sydenham chorea is present, the onset of which might be very delayed.(3) The small number of definite cases who lacked evidence of streptococcal infection on serology or throat swab may have had chorea, may have had serological tests too early in disease without opportunity for convalescent serology to be done, or the clinician may have chosen to override the recommended diagnostic criteria.

The cumulative incidence risk of disease progression per year for those diagnosed with possible ARF stopped increasing at 11%, 4 years from initial diagnosis, and for those diagnosed with probable ARF at 14%, 3 years from initial diagnosis. In contrast, the cumulative incidence risk of disease progression after definite ARF continued to increase each year since initial diagnosis, from 9% at 1 year to 34% at 5.5 years from initial diagnosis (Table 3). While the trend of increasing risk of disease progression from initial diagnosis largely concurs with previous studies, the cumulative incidence risk of progression to RHD after definite ARF was lower in our study. For example, a study of 1997-2013 Northern Territory data found that the cumulative risk of progression from definite ARF to RHD increased from 27% at 1 year from initial diagnosis to 52% at 10 years since initial diagnosis,(9) and another study of 1997-2010 data reported that the risk of progression from definite ARF to RHD was 35% at 1 year and 61% at 10 years.(19) The lower rate of disease progression in our study may reflect exclusion of the most-severe ARF cases from cumulative incidence calculations, since they already had the outcome of interest (RHD) at baseline. Other explanations may include improving delivery of secondary prophylaxis which occurred during our study period(20), or greater clinical detection over time of more subtle ARF cases which are less likely to progress, which the increasing ARF detection rates reported here suggest.

Cox proportional hazards analysis demonstrated that those with more certain ARF were more likely to experience disease progression (compared to those with possible ARF). The risk of progression from no RHD to RHD was significantly higher if the initial diagnosis was definite ARF compared to possible ARF (p<0.001 for both mild and moderate-severe RHD outcomes) (Table 6). Those who had only joint manifestations at initial diagnosis of ARF were 52% less likely to experience disease progression (compared to carditis and/or chorea). Joint-only manifestations, which may only fulfill criteria for possible or probable ARF, may in other settings be labelled as ‘post streptococcal reactive arthritis’, considered to be a separate entity from ARF.(21, 22) However, since 6.6% (37/560) of people still developed RHD in our series after initial joint-only presentations, we consider a diagnosis of ARF and provision of secondary prophylaxis to be safer in our context.

In this dataset, people with ARF who had greater penicillin adherence were also those with a higher risk of disease progression - but this association was not statistically significant (Table 4). Furthermore, of the cases that progressed (and had adherence data), less than half (40/86; 47%) were adherent to ≥80% of their BPG injections and adherence for both definite and probable ARF cases was shown to decline over time from initial diagnosis (Figure 3). These findings may be attributable to the association between disease severity and adherence. Previous research using the same NT register, but an earlier time period, identified that those with most severe disease (e.g. requiring surgery) were those who were subsequently most adherent, potentially attributable to higher-level health system supports implemented for these high-risk individuals, and greater patient understanding of the risks of non-adherence. (10) The same paper showed a clear protective effect of adherence on ARF outcomes when adherence prior to the event of interest was examined. Therefore, we conclude that adherence reduces ARF recurrences or RDH progression, but in this dataset, we were unable to demonstrate this relationship. Also, we used percent adherence which does not necessarily factor in ‘days at risk’(10); that is, the timing of dose administration (e.g. 13 BPG doses may be given in a 12 month period, but if there are gaps of greater than 28 days, then the patient will be at risk of ARF recurrence during those times).

Strengths of this study include that it is the first to include outcomes for possible and probable ARF cases and that ARF is notifiable in the NT and therefore ARF diagnoses are well captured and documented in the NT RHD Register. A limitation of this study is that a longer period of follow-up time may have captured more events. For example, disease progression from ARF to RHD is known to still occur at 10 years from initial diagnosis.(19) However, for possible and probable ARF, the risk of disease progression appeared to stabilise approximately 4 years from initial diagnosis (within the 5.5-year study timeframe). Another limitation is missed diagnoses of ARF; up to 75% of newly diagnosed cases of RHD in northern Australia have not been previously diagnosed with ARF.(3, 23) As noted, concurrent ARF and RHD diagnoses were excluded since they had a disease-free survival time of zero so could not be included in analysis of progression. Exclusion of these most severe ARF cases impacted sample size and statistical rigour, follow-up time and the ability to calculate median survival time, an important factor in the interpretation of survival data.(24)

## Conclusion

This study provides much needed data about the likelihood of disease progression for children and young people given a diagnosis of possible and probable ARF. Children and their families can be reassured that possible ARF, probable ARF and ARF affecting the joints only, carry relatively good prognoses. These diagnoses are more consistent with post-streptococcal reactive arthritis than true ARF. However, secondary prophylaxis with penicillin is still of critical importance given the morbidity of recurrences and the small but present risk of progression to RHD. The data provide good news that clinicians appear to be detecting and reporting more cases of uncertain ARF over time, a critical step in avoiding missed opportunities for prevention.

## Data Availability

The raw data underlying the results presented in the study are available from the Northern Territory Rheumatic Heart Disease Register, contactable through the NT Centre for Disease Control: https://nt.gov.au/wellbeing/health-conditions-treatments/rheumatic-heart-disease.

## Acknowledgements

This project was undertaken as part of the Master of Public Health at the University of Melbourne. The authors would like to thank Dr Jessica de Dassel, RHD Registry Data Analyst, and Dr Vicki Krause, Northern Territory Centre for Disease Control Director, for facilitating the data request and approval.

